# A dataset of multi-contrast unbiased average MRI templates of a Parkinson’s disease population

**DOI:** 10.1101/2022.07.06.22277331

**Authors:** Victoria Madge, Vladimir S Fonov, Yiming Xiao, Lucy Zou, Courtney Jackson, Ronald B Postuma, Alain Dagher, Edward A Fon, D Louis Collins

## Abstract

Parkinson’s disease (PD) is a complex neurodegenerative disorder affecting regions such as the substantia nigra (SN), red nucleus (RN) and locus coeruleus (LC). Processing MRI data from patients with PD requires anatomical structural references for spatial normalization and structural segmentation. Extending our previous work [1][2], we present multi-contrast unbiased MRI templates using nine 3T MRI modalities: T1w, T2*w, T1-T2* fusion, R2*, T2w, PDw, fluid-attenuated inversion recovery (FLAIR), improved susceptibility-weighted imaging (CLEAR-SWI) [3], and neuromelanin-sensitive MRI (NM). Using our previous methods to build unbiased average templates [4], 1 mm isotropic voxel size templates were created, along with 0.5 mm isotropic whole brain templates and 0.3 mm isotropic templates of the midbrain. All templates were created from 126 PD patients (44 female; ages=40-87), and 17 healthy controls (13 female; ages=39-84), except the NM template, which was created from 85 PD patients and 13 controls, respectively. The dataset is available on the NIST MNI Repository via the following link: http://nist.mni.mcgill.ca/multi-contrast-pd126-and-ctrl17-templates/.

## Specifications table

|  |  |
| --- | --- |
| **Subject** | Health and medical sciences |
| **Specific subject area** | Brain MRI templates of Parkinson's disease population |
| **Type of data** | Images, figures, volume data |
| **How the data were acquired** | 3T Magnetic resonance imaging (Siemens 3T Prisma MRI scanner) |
| **Data format** | Processed |
| **Description of data collection** | Subjects were recruited through the Quebec Parkinson's Network (QPN), PD patients with a PD diagnosis, inclusion and exclusion criteria based on [5], and age- and sex- matched healthy controls. 3T MRI sequences used to make templates include: T1w MPRAGE, multi-echo GRE [1], T2w PD, 2D FLAIR, T1-weighted Turbo Spin Echo (NM) [6]. Raw images went through denoising [7], non-uniformity correction [8], and intensity normalization. |
| **Data source location** | Institution: Montreal Neurological Institute (MNI)<br>City/Town/Region: Montreal<br>Country: Canada |
| **Data accessibility** | The dataset is available on the NIST MNI Repository via the following link: http://nist.mni.mcgill.ca/multi-contrast-pd126-and-ctrl17-templates/. |

## Value of the data

- These publicly available templates were created using 126 PD patients while current templates use much smaller cohorts.
- Accompanying templates of 17 age-matched healthy controls facilitates group-wise analyses.
- Templates represent averaged anatomical and MRI intensity features of Parkinson’s disease, and healthy controls, in nine different MRI contrasts, respectively.

## Data description

The dataset is a collection of nine multi-contrast brain MRI templates, created from 3T scans of 126 Parkinson’s disease patients, with an accompanying set of nine templates of the same MRI contrasts created from 17 healthy controls. The templates are in ICBM152 stereotaxic space, in three different resolutions: 1 × 1 × 1 mm^3^, 0.5 × 0.5 × 0.5 mm^3^, and 0.3 × 0.3 × 0.3 mm^3^.

**Figure 1:**
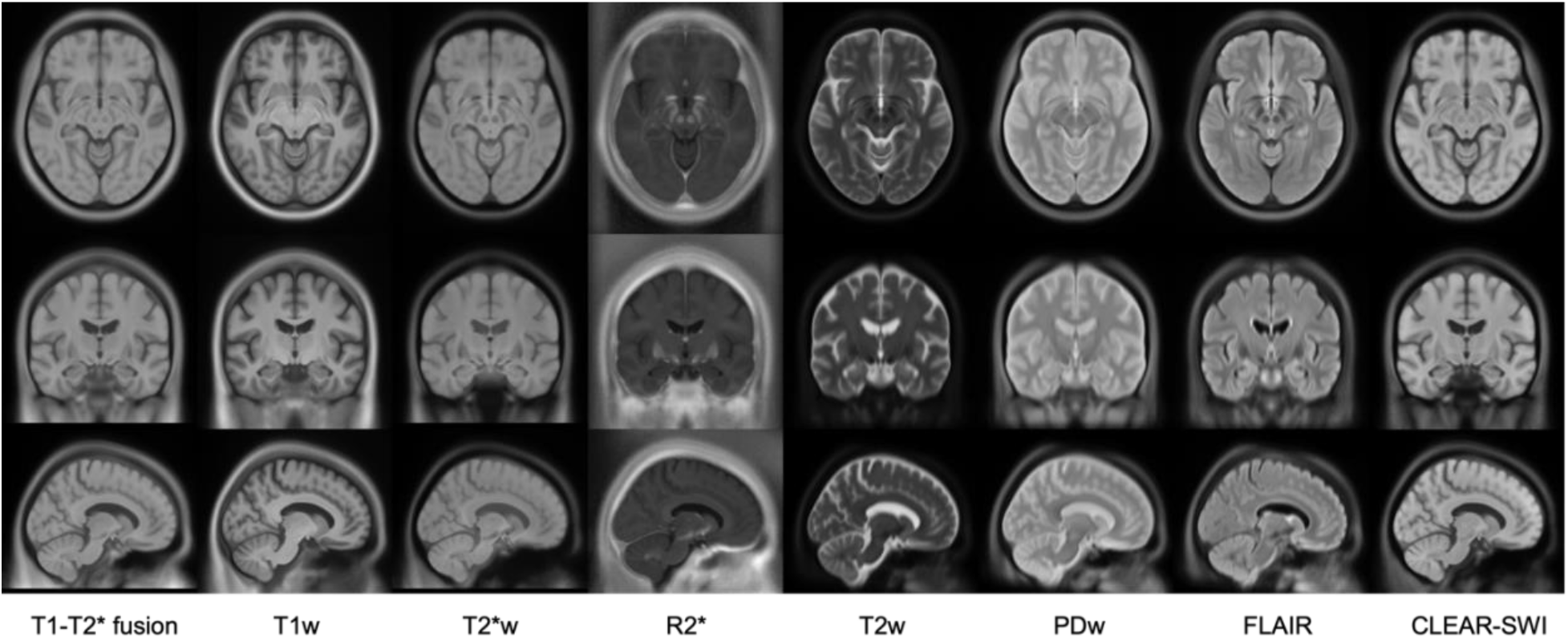
Multi-contrast population of 126 PD patients (voxel size 1 × 1 × 1 mm^3^), showing eight contrasts in columns from left to right: T1-T2* fusion, T1w, T2*w, R2*, T2w, PDw, FLAIR, CLEAR-SWI. The templates are shown in three slices: axial (row 1), coronal (row 2), and sagittal (row 3).

**Figure 2:**
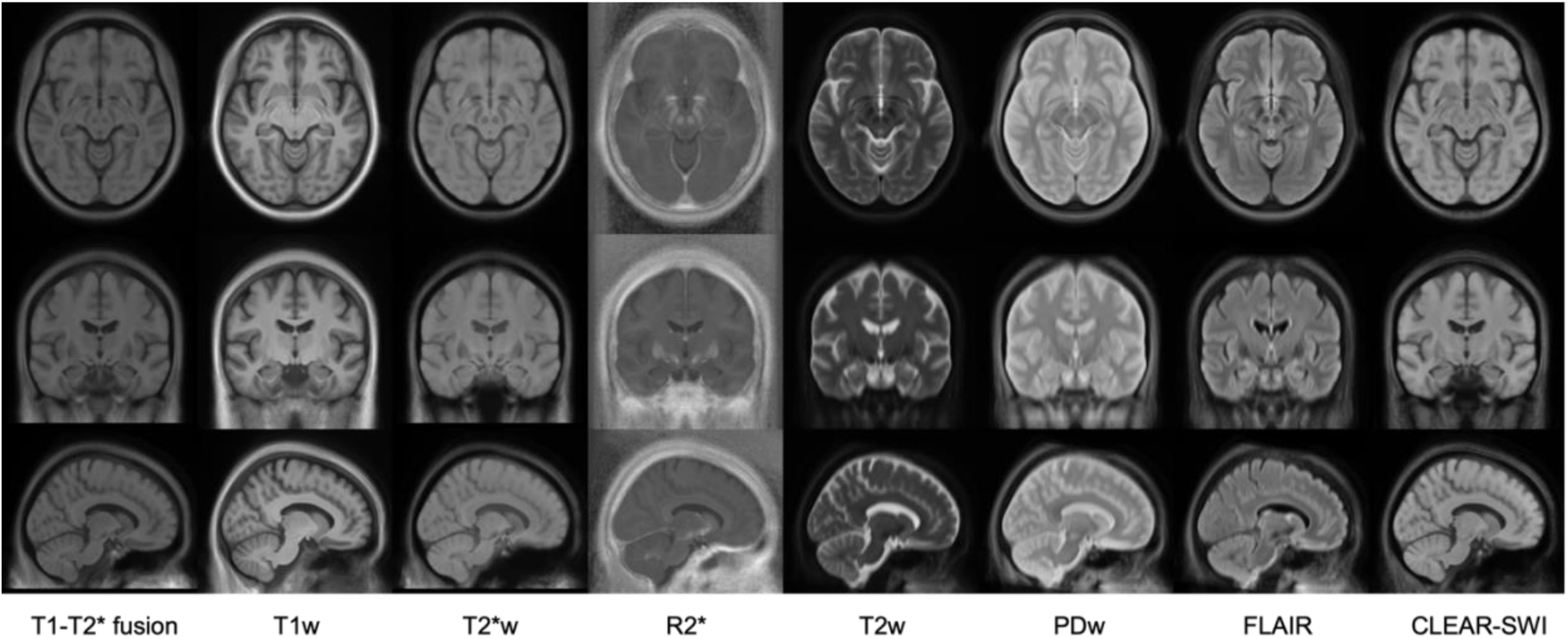
Multi-contrast population of 17 healthy controls (voxel size 1 × 1 × 1 mm^3^), showing eight contrasts in columns from left to right: T1-T2* fusion, T1w, T2*w, R2*, T2w, PDw, FLAIR, CLEAR-SWI. The templates are shown in three slices: axial (row 1), coronal (row 2), and sagittal (row 3).

**Figure 3:**
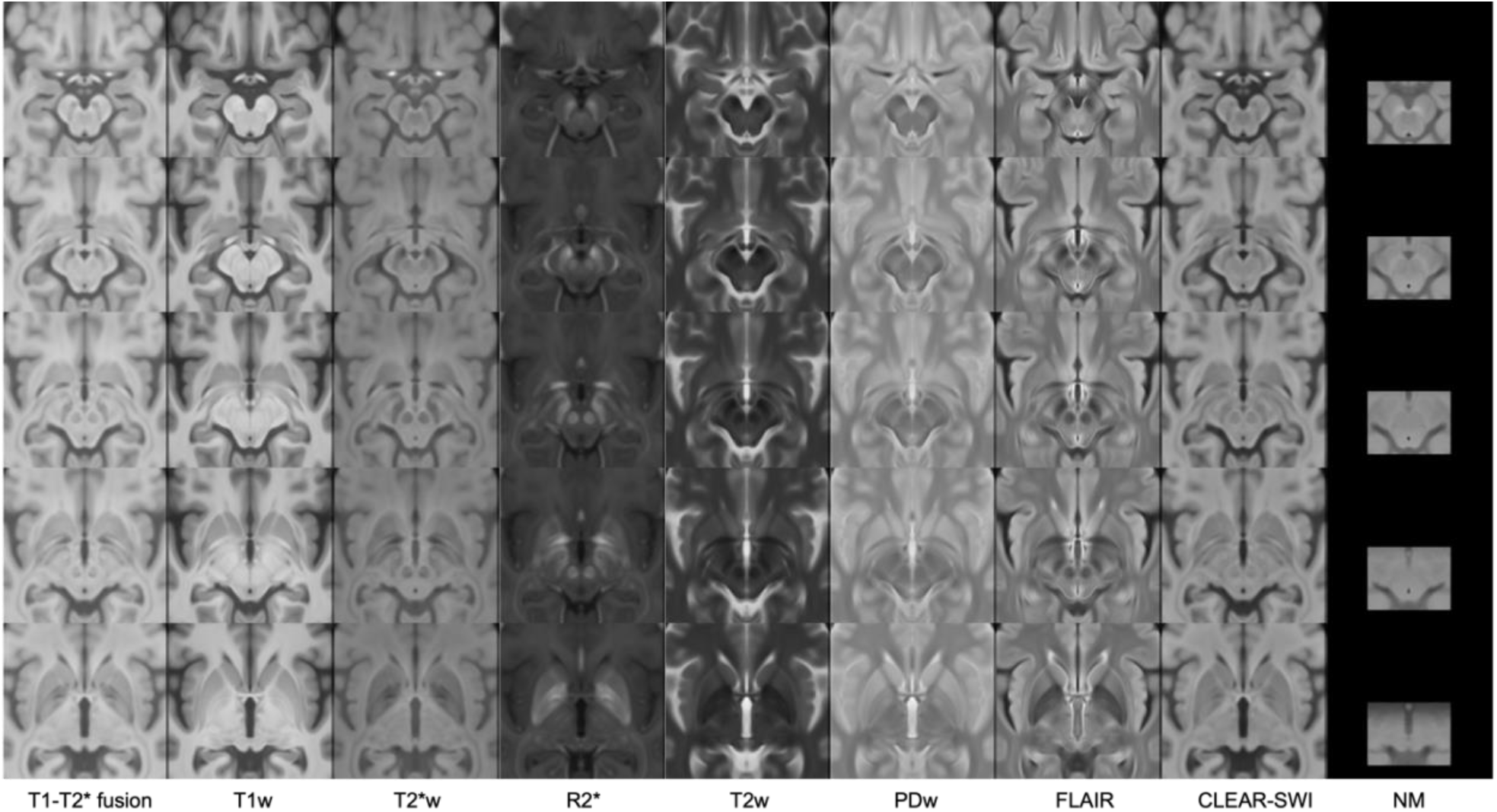
Multi-contrast population of 126 PD patients (voxel size 0.3 × 0.3 × 0.3 mm^3^), showing nine contrasts in columns from left to right: T1-T2* fusion, T1w, T2*w, R2*, T2w, PDw, FLAIR, CLEAR-SWI, NM. The templates are shown as five axial slices from superior to inferior direction (rows from top to bottom at z= 87,101,108,115,129 mm in stereotaxic space). Note that the NM template is derived from 85 patients only, and from an NM-sensitive sequence which is not 3D and does not cover the full extent of the brain.

**Figure 4:**
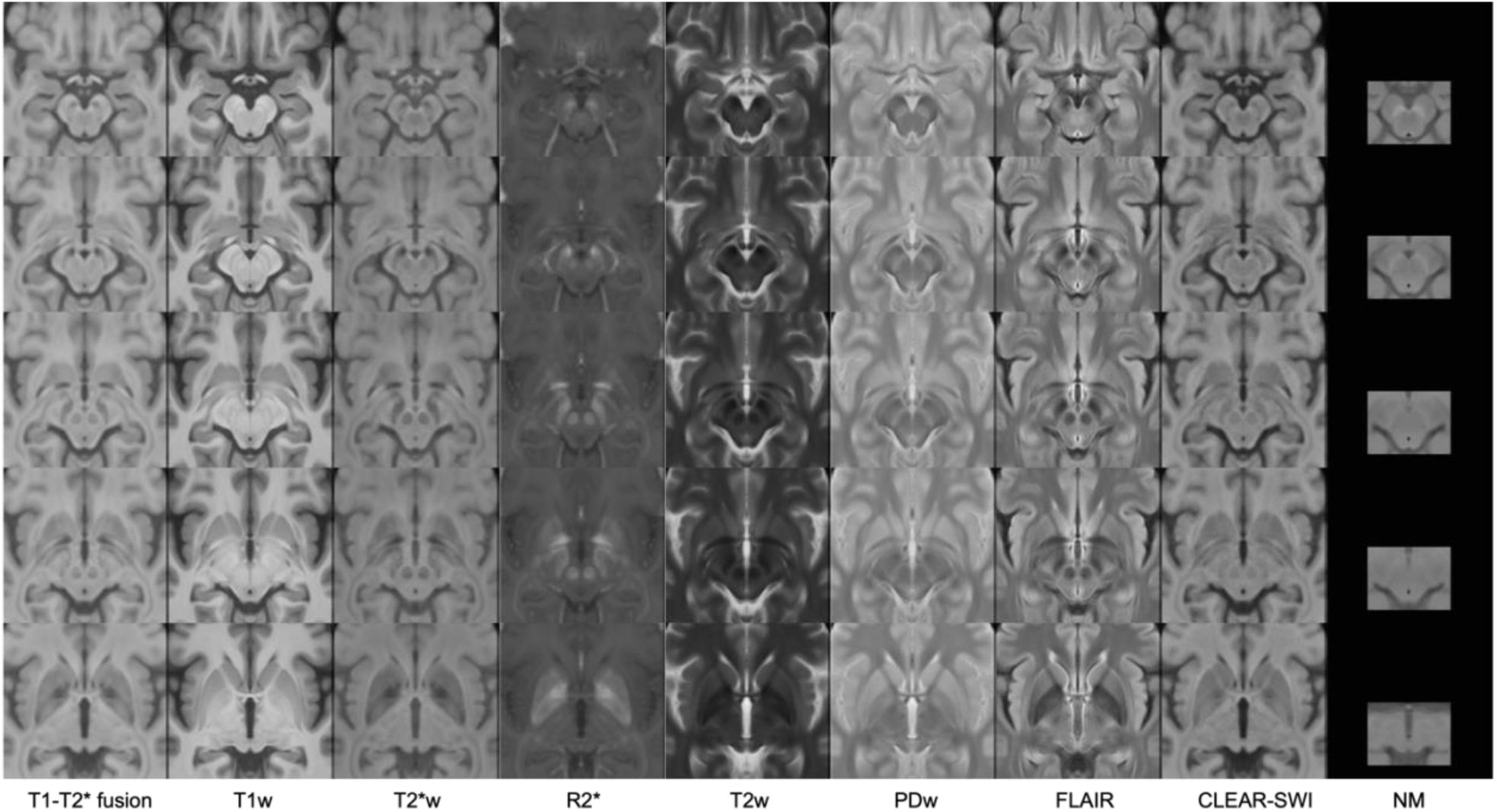
Multi-contrast population of 17 healthy controls (voxel size 0.3 × 0.3 × 0.3 mm^3^), showing nine contrasts in columns from left to right: T1-T2* fusion, T1w, T2*w, R2*, T2w, PDw, FLAIR, CLEAR-SWI, NM. The templates are shown as five axial slices from superior to inferior direction (rows from top to bottom at z= 87,101,108,115,129 mm in stereotaxic space). Note that the NM template is derived from 13 subjects only, and from an NM-sensitive sequence which is not 3D and does not cover the full extent of the brain.

## Experimental design, materials and methods

### Subjects

All Parkinson’s patients and healthy controls were recruited through the registry of the Quebec Parkinson Network from 2018 onward and are included in the Montreal Neurological Institute’s (MNI) Open Science Clinical Biospecimen Imaging and Genetic (C-BIG) Repository [9]. After rigorous and detailed informed consent, 126 PD patients and 17 controls were scanned, and their age, sex, Unified Parkinson’s Disease Rating Scale Part III (UPDRS-III) scores, and Hoen and Yahr (H&Y) scores were recorded. Table 1 shows the demographics of each of the population cohorts.

| Disease status | Sex (male/female) | Age | UPDRS-III score | H&Y score |
| --- | --- | --- | --- | --- |
| PD | 82 / 44 | 64.8 ± 9.1 | 29.9 ± 15.3* | 1.8 ± 0.68** |
| controls | 4 / 13 | 64.0 ± 11.4 | n/a | n/a |

\*UPDRS-III scores are calculated for 118 PD patients only (those that were available)
\*\*H&Y scores are calculated for 66 patients only (those that were available)

### Image acquisition

Each subject was scanned on a Siemens 3T Prisma magnetic resonance imaging (MRI) scanner with the following sequences and accompanying parameters listed in Table 2. From the multi-echo GRE data, several imaging contrasts were created: T2*w image, R2* (i.e., 1/T2*), and CLEAR-SWI [3]. All image contrasts were acquired simultaneously at the same imaging session, and are thus inherently co-registered, per subject.

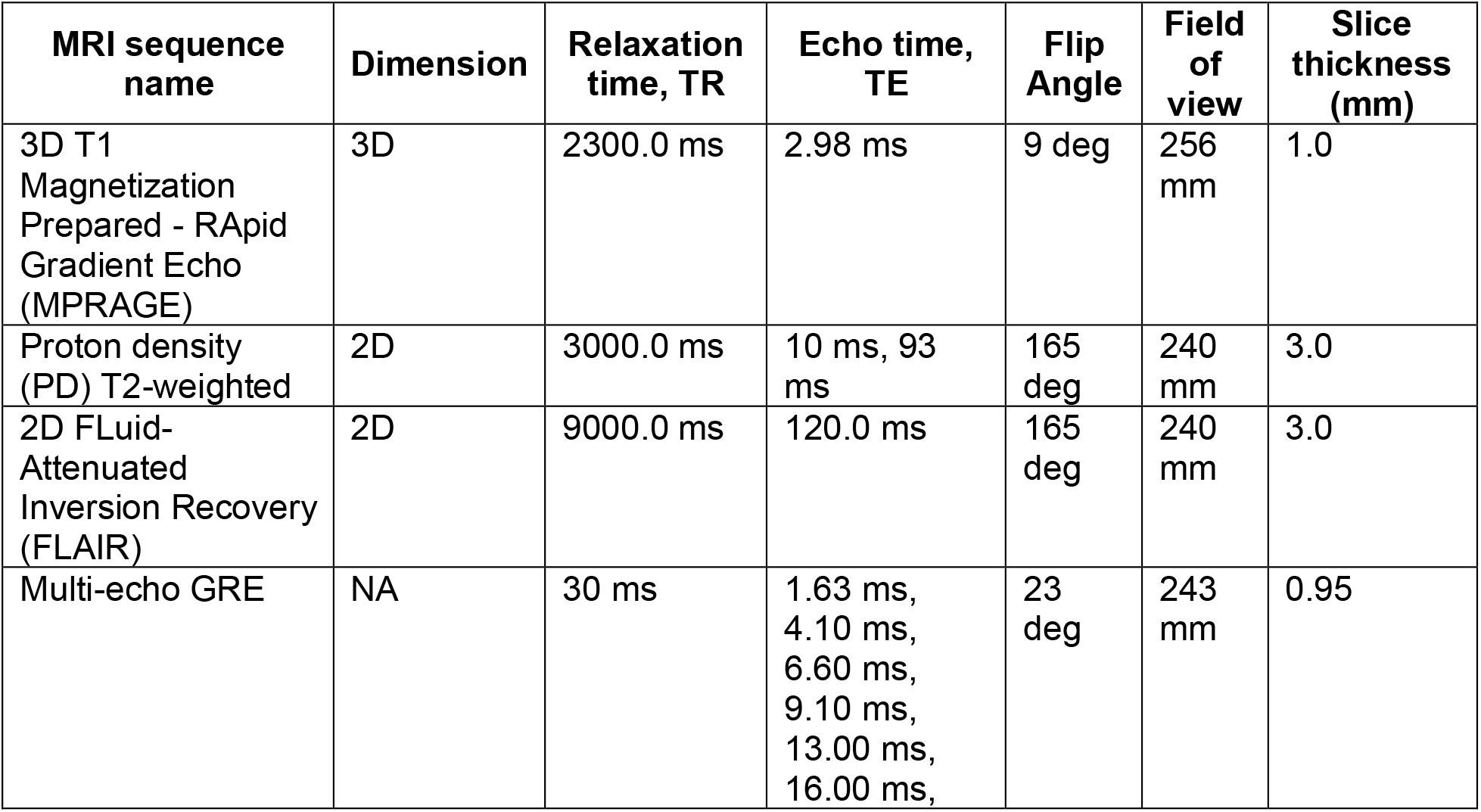
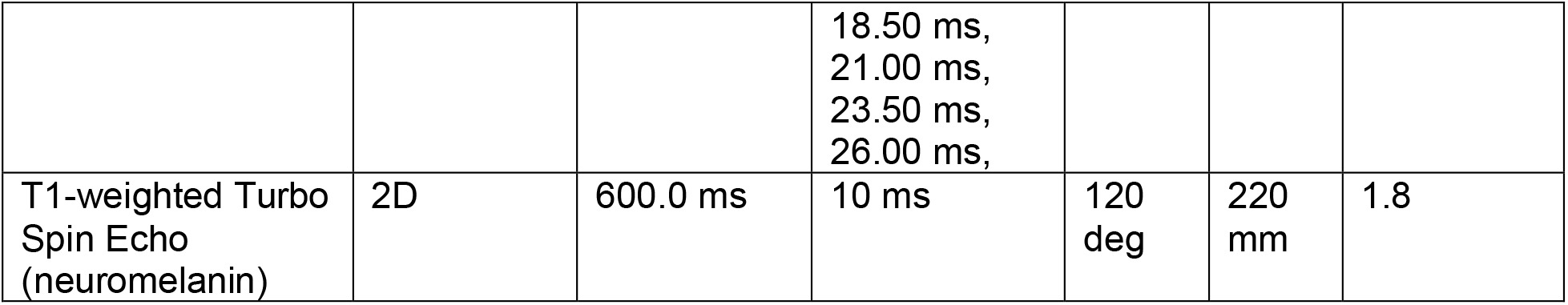

### Image processing and template creation

For each of the imaging modalities, brain masks were generated using BEaST: brain extraction based on nonlocal segmentation technique [10] using the T1w image. After non-local means denoising [7], non-uniformity correction [8], and intensity normalization, each image was registered to stereotaxic space using an in-house PPMI template (which is in ICBM152 stereotaxic space) as the stereotaxic registration target [4].

T1-T2* fusion MR images were created for each subject following methods from [11]. Briefly, T1w and T2*w images from the same subject are added together, using spatially varying weights calculated from the R2* volume (i.e., 1/T2*), such that the T1w structure near the cortex and T2*w deep grey nuclei are preserved. Improved susceptibility-weighted images (CLEAR-SWI) were created from methods described in [3]. All other imaging modalities (T1, T2*w, R2*, T2w, PDw, FLAIR, and NM) were ready for template creation.

The T1-T2* fusion images were used to create the transformations that would drive the unbiased average template creation for all imaging modalities. Template creation follows the methods from [4], where hierarchical registration using Automatic Nonlinear Image Matching and Anatomical labelling (ANIMAL) [12] is employed to register an initial template model (here, the MNI PD25 template [1] [2] was used) to each of the subjects. This process is performed iteratively, correcting for residual mis-registration at each step, until convergence is reached. Once convergence is reached and the unbiased average T1-T2* fusion template is created, the individual subject-to-template transformations are saved and used to create the templates for the remaining contrasts (T1, T2*w, R2*, T2w, PDw, FLAIR, NM, and CLEAR-SWI). The same transformations were used to create 0.5 mm and 0.3 mm templates, resampling the pre-processed MRI volumes to the appropriate resolution in stereotaxic space.

## Ethics statements

The MR imaging and motor evaluation data collected and shared through the MNI’s Open Science C-BIG repository [9] are collected from patients, recruited through the registry of the Quebec Parkinson Network, with a rigorous and detailed informed consent. The de-identified imaging and motor data are collected under protocols that comply with the Health Insurance Portability and Accountability Act of 1996 (HIPAA), the Declaration of Helsinki, and the MUHC Research Ethics Board (C-BIG general protocol: 2017-330, 15-944-MUHC; C-BIG imaging protocol: 2019-4759; QPN protocol: 2015-143, MP-CUSM-NEU-14-053, MP-37-2015-143).

## Data Availability

All data produced are available online at: http://nist.mni.mcgill.ca/multi-contrast-pd126-and-ctrl17-templates/

## CRediT author statement

Victoria Madge: Conceptualization, Methodology, Software, Writing – Original Draft, Visualization

Vladimir S Fonov: Methodology, Software, Data Curation, Writing – Review & Editing

Yiming Xiao: Methodology, Software, Writing – Review & Editing

Lucy Zou: Resources

Courtney Jackson: Resources

Ronald B Postuma: Supervision

Alain Dagher: Conceptualization, Supervision, Project administration

Edward A Fon: Project administration

D Louis Collins: Conceptualization, Methodology, Software, Writing – Review & Editing, Supervision, Project administration, Funding acquisition

## Acknowledgments

This study was supported by the Weston Grant [RR171117].

## References

[1] Y. Xiao et al., “Multi-contrast unbiased MRI atlas of a Parkinson’s disease population,” Int. J. Comput. Assist. Radiol. Surg., vol. 10, no. 3, pp. 329–341, Mar. 2015.

[2] Y. Xiao et al., “A dataset of multi-contrast population-averaged brain MRI atlases of a Parkinson?s disease cohort,” Data Br., vol. 12, pp. 370–379, Jun. 2017.

[3] K. Eckstein et al., “Improved susceptibility weighted imaging at ultra-high field using bipolar multi-echo acquisition and optimized image processing: CLEAR-SWI,” Neuroimage, vol. 237, p. 118175, Aug. 2021.

[4] V. Fonov, A. C. Evans, K. Botteron, C. R. Almli, R. C. McKinstry, and D. L. Collins, “Unbiased average age-appropriate atlases for pediatric studies,” Neuroimage, vol. 54, no. 1, pp. 313–327, Jan. 2011.

[5] NICE, “Parkinson’s Disease [CG35]: National clinical guideline for diagnosis and management in primary and secondary care,” R. Coll. Physicians, London, 2006.

[6] M. Sasaki et al., “Neuromelanin magnetic resonance imaging of locus ceruleus and substantia nigra in Parkinson’s disease,” Neuroreport, vol. 17, no. 11, pp. 1215–1218, Jul. 2006.

[7] P. Coupe, P. Yger, S. Prima, P. Hellier, C. Kervrann, and C. Barillot, “An optimized blockwise nonlocal means denoising filter for 3-D magnetic resonance images,” IEEE Trans. Med. Imaging, vol. 27, no. 4, pp. 425–441, Apr. 2008.

[8] J. G. Sled, A. P. Zijdenbos, and A. C. Evans, “A nonparametric method for automatic correction of intensity nonuniformity in mri data,” IEEE Trans. Med. Imaging, vol. 17, no. 1, pp. 87–97, 1998.

[9] S. Das et al., “The C-BIG Repository: an Institution-Level Open Science Platform,” Neuroinformatics, 2021.

[10] S. F. Eskildsen et al., “BEaST: Brain extraction based on nonlocal segmentation technique,” Neuroimage, vol. 59, no. 3, pp. 2362–2373, Feb. 2012.

[11] Y. Xiao et al., “Patch-based label fusion segmentation of brainstem structures with dual-contrast MRI for Parkinson’s disease,” Int. J. Comput. Assist. Radiol. Surg., vol. 10, no. 7, pp. 1029–1041, 2015.

[12] D. L. Collins and A. C. Evans, “ANIMAL: validation and applications of non-linear registration-based segmentation,” Int. J. Pattern Recognit. Artif. Intell., vol. 11, no. 8, pp. 94–116, 1997.

